## Supplementary figures and images for "Estimating pulse wave velocity from the radial pressure wave using machine learning algorithms"

### Supplemental Figure 1

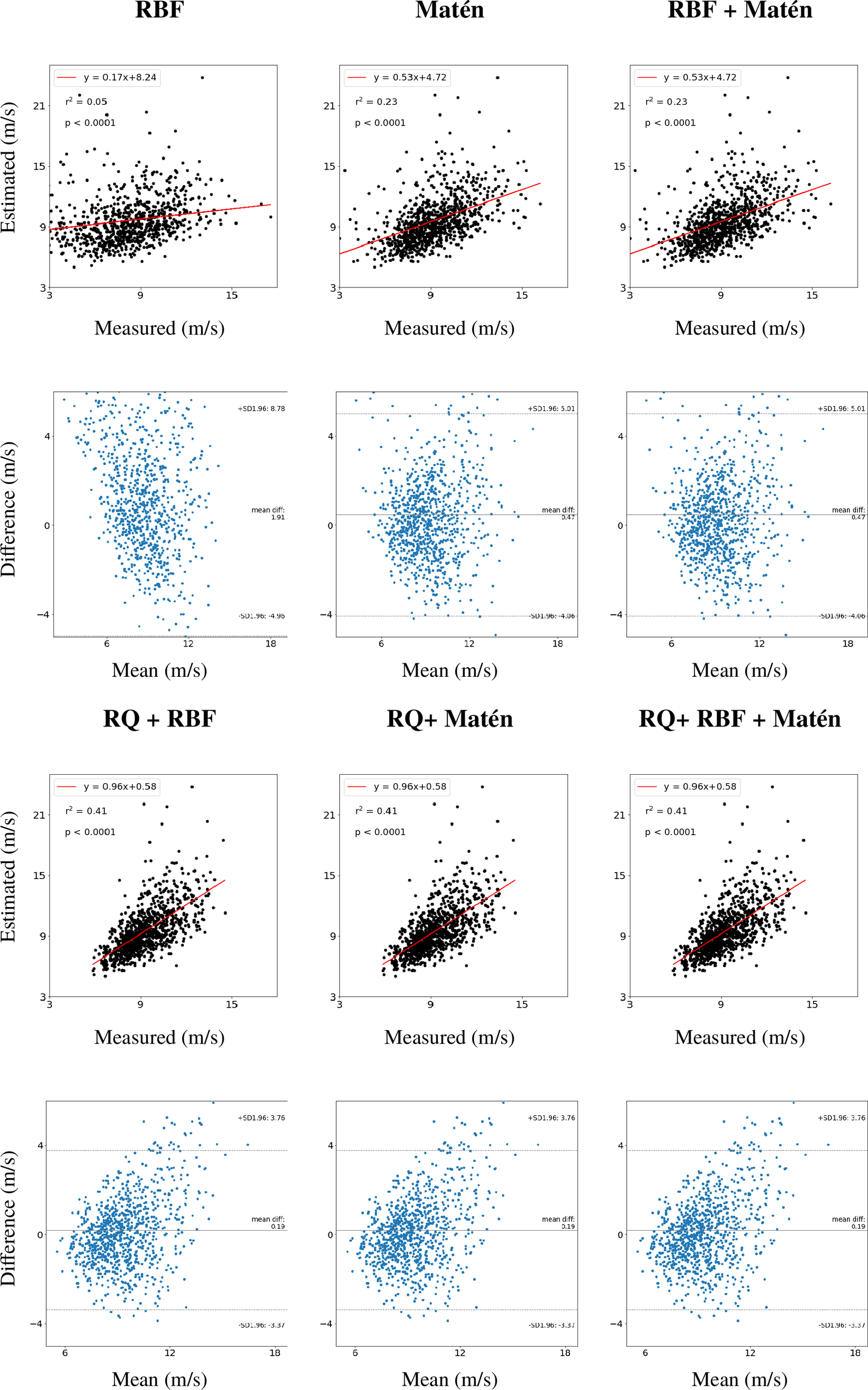

### Supplemental Figure 2

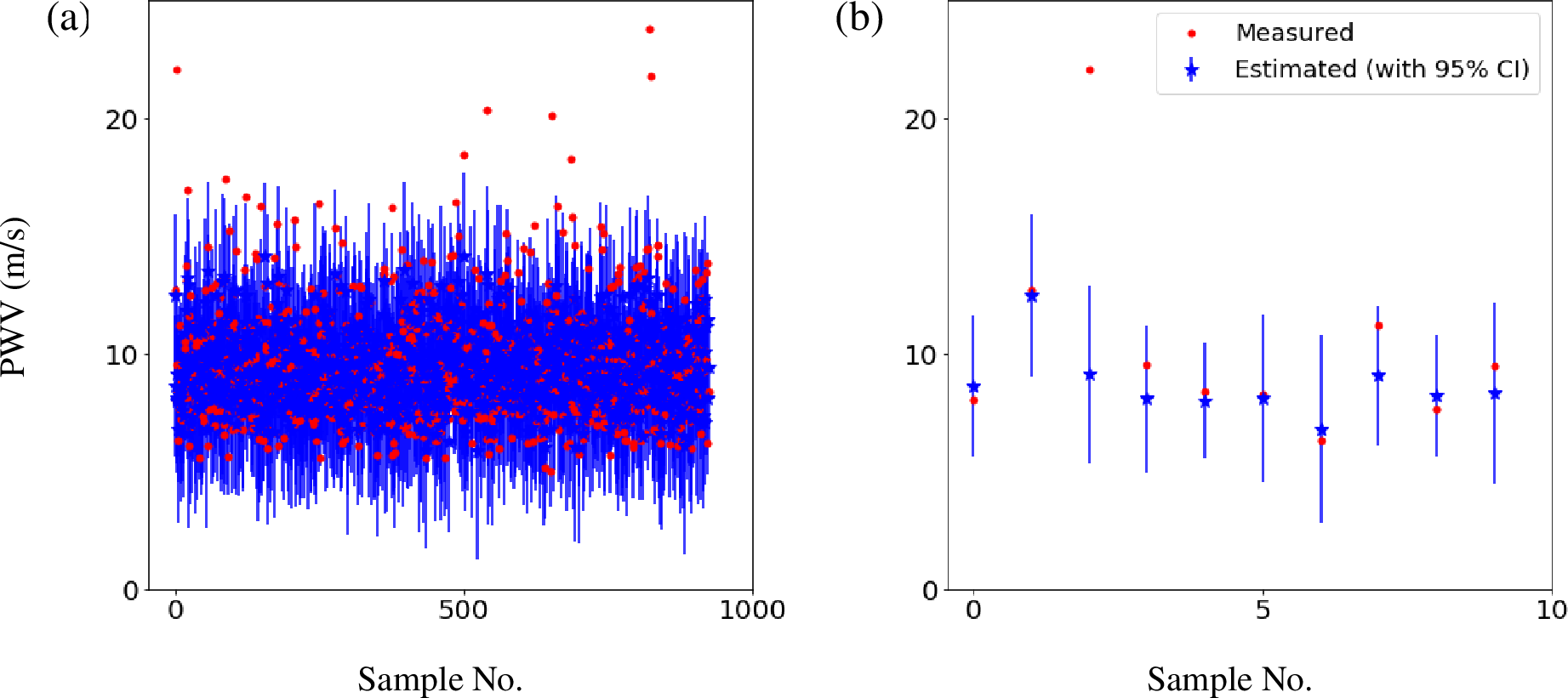

### Supplemental Figure 3

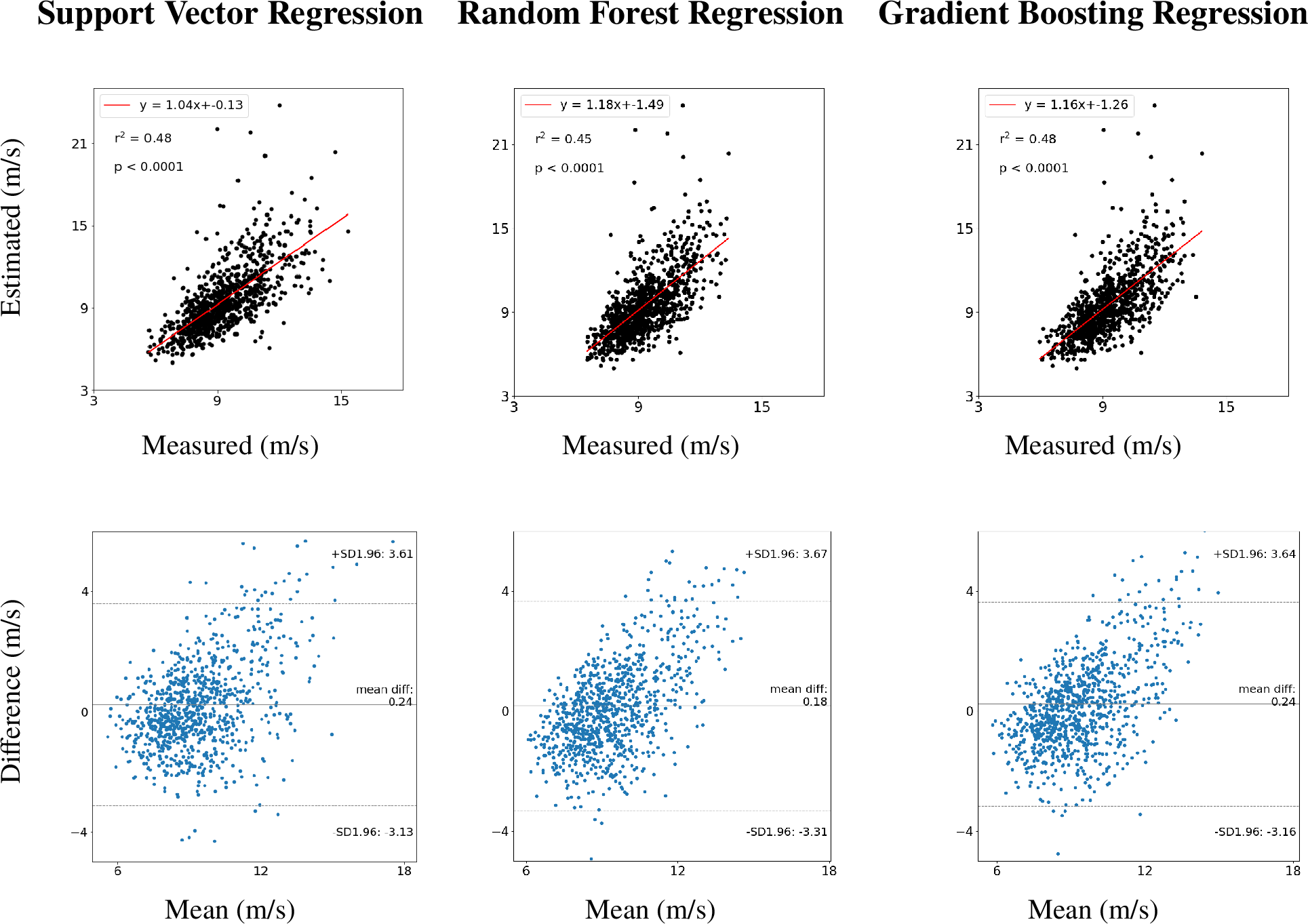

### Supplemental Figure 4

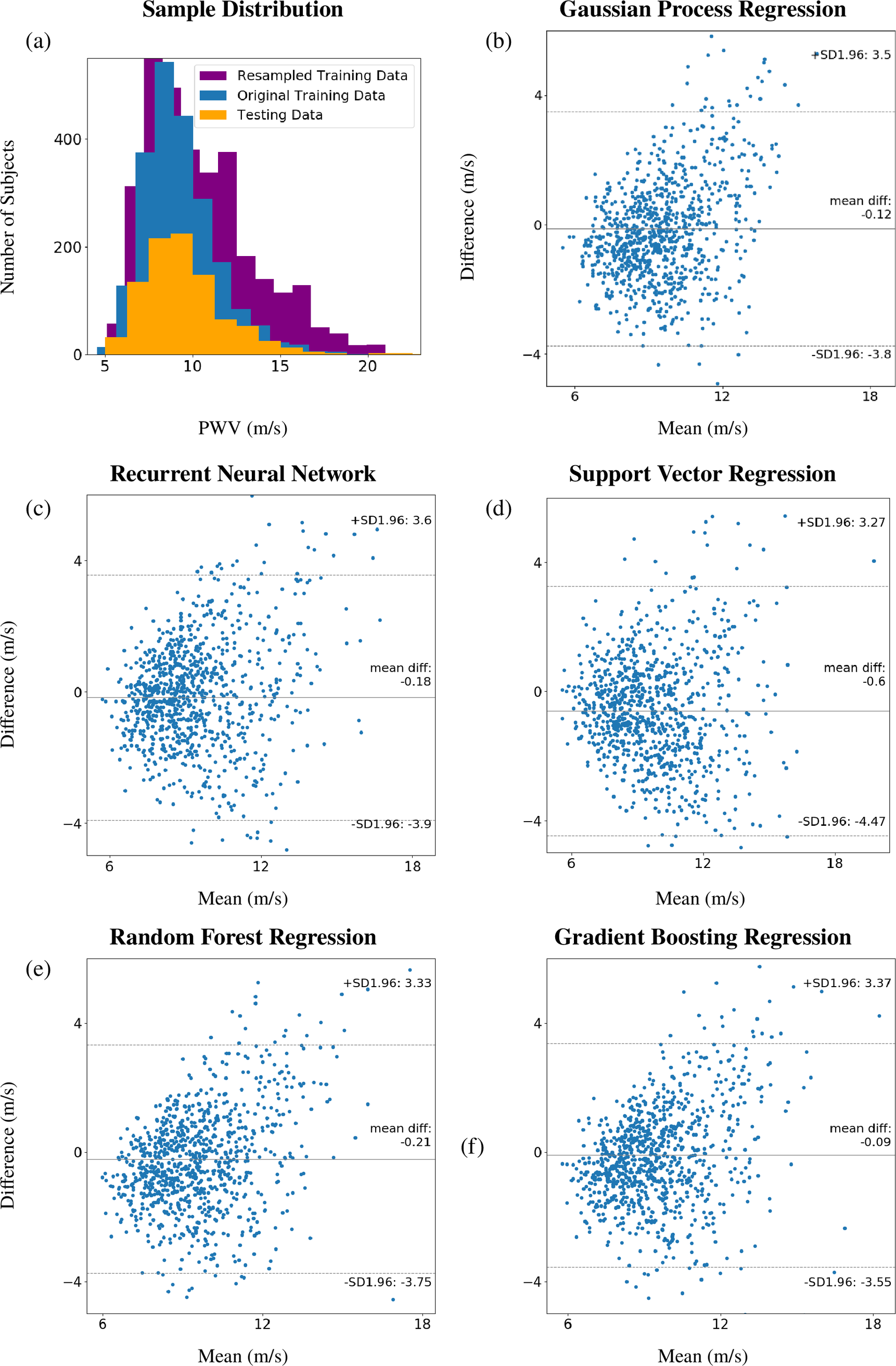

### Supplemental Figure 5

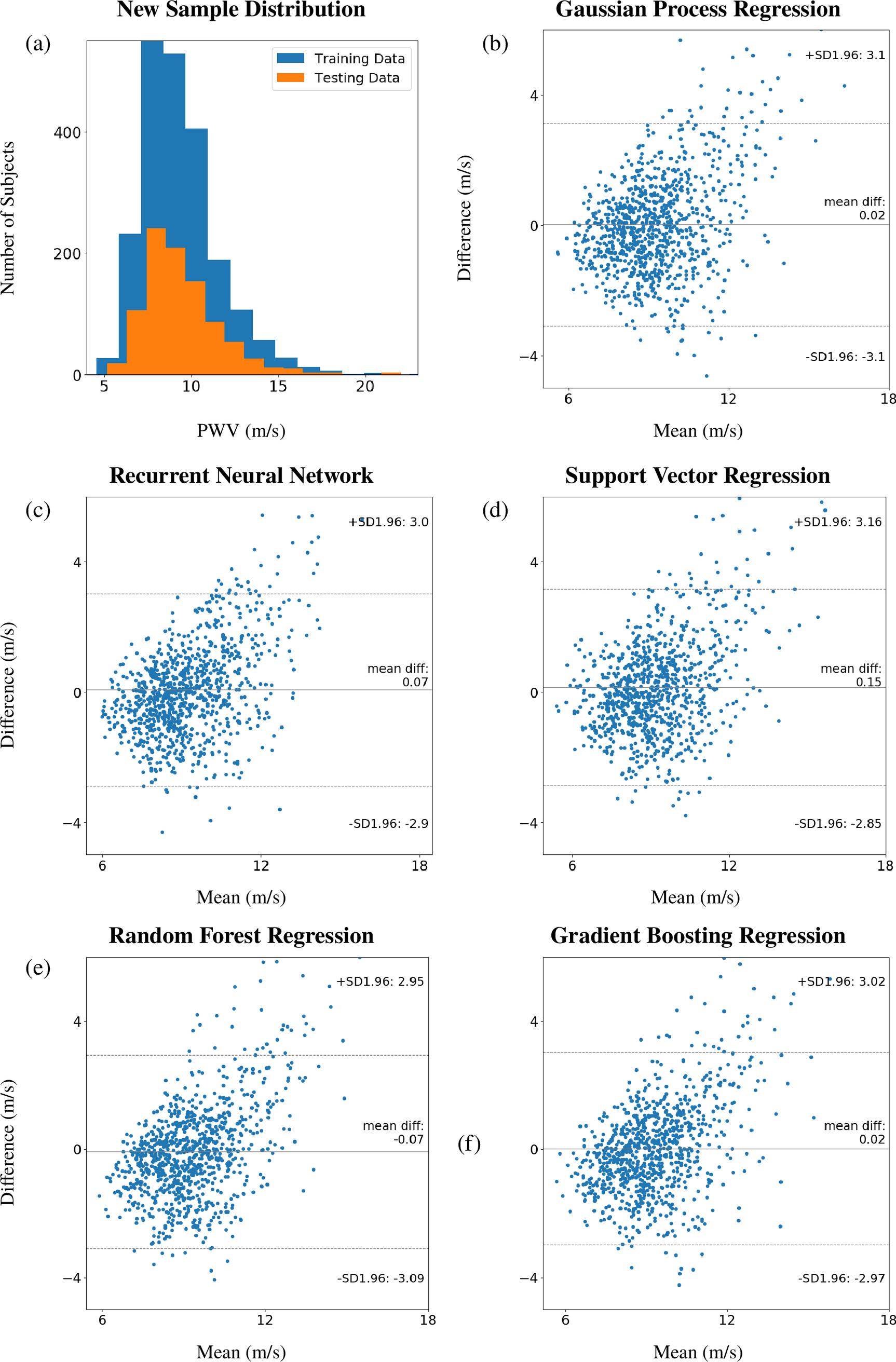
